# Optical Genome Mapping And Single Nucleotide Polymorphism Microarray: An Integrated Approach For Investigating Challenging Cases Of Products Of Conception

**DOI:** 10.1101/2022.02.03.22269494

**Authors:** Nikhil S Sahajpal, Ashis K Mondal, Sudha Ananth, Chetan Pundkar, Kimya Jones, Colin Williams, Timothy Fee, Amanda Weissman, Giuseppe Tripodi, Eesha Oza, Larisa Gavrilova-Jordan, Nivin Omar, Alex Hastie, Barbara R DuPont, Lawrence Layman, Alka Chaubey, Ravindra Kolhe

## Abstract

Conventional cytogenetic analysis of products of conception (POC) is of limited utility because of failed cultures, microbial and maternal cell contamination (MCC). Optical genome mapping (OGM) is an emerging technology that has the potential to replace conventional cytogenetic methods. The use of OGM precludes the requirement for culturing (and related microbial contamination). However, a high percentage of MCC impedes a definitive diagnosis, which can be addressed by an additional pre-analytical quality control step that includes histological assessment of H&E stained slides from FFPE tissue with macro-dissection for Chorionic villi to enrich fetal tissue component for Single nucleotide polymorphism (SNP) microarray analysis. An internal audit of POC cases subjected to karyotype-only analysis showed a low yield on clinically actionable information that contributed to patient care. To improve the diagnostic yield, an integrated workflow was devised that included MCC characterization of POC tissue, followed by OGM for MCC negative cases or SNPM with histological assessment for MCC positive cases. A result was obtained in 93% (29/31) cases with a diagnostic yield of 45.1% (14/31) with proposed workflow compared to 9.6% (3/31) and 6.4% (2/31) with routine workflow, respectively. The integrated workflow with these technologies demonstrates the clinical utility and higher diagnostic yield in evaluating POC specimens.

## Introduction

Clinically recognized pregnancy losses (at any gestational age) occur in ∼15-25% of pregnancies and is traumatic event for women and their families.^1^ The rate of miscarriage (pregnancy loss <☐20☐weeks’ gestation) is reported to be ∼15-20%,^2^ while 1/100 pregnancies result in stillbirth (pregnancy loss ≥☐20☐weeks) in the US.^3^ The majority of miscarriages are spontaneous and a significant number result from genetic anomalies influenced by maternal age.^1,4,5^ The rate of miscarriage increases with age as women with ≤ 35 years reported a 9-12% rate of miscarriage in 6-12 weeks;^6,7^ > 35 years of age had a high incidence of trisomic pregnancies,^4^ and those > 40 years of age had an ∼50% rate of miscarriage.^1,7,8^ In contrast, recurrent pregnancy loss is a disorder defined as two or more failed clinical pregnancies. The standard screening protocol as recommended by the American Society for Reproductive Medicine (ASRM) comprises determining parental karyotypes, antiphospholipid antibodies, uterine cavity evaluation, thyrotropin, and prolactin levels, after two consecutive failed clinical pregnancies. Chromosomal analysis of products of conception (POC) has been deemed useful in the setting of ongoing therapy for recurrent pregnancy loss (RPL).^9^

Chromosomal abnormalities provide a genetic diagnosis that is critical in understanding the cause of miscarriage and for recurrent-risk counseling that might help identify familial chromosomal rearrangements that predispose the parents to these events or the birth of children with genetic defects. Most pregnancy losses are marked with aneuploidies detected through traditional cytogenetic techniques.^10,11^ However, the techniques are limited by the requirement of procuring live dividing cells, possible microbial or maternal cell contamination (MCC), and poor chromosome morphology that preclude a definitive diagnosis. The challenges associated with the cytogenetic analysis of POC demonstrate the need for alternate technologies or methodologies to improve the diagnostic yield in this important area of reproductive medicine.

Optical genome mapping (OGM) is an emerging technology that has demonstrated the potential to replace conventional cytogenetic methods in several recent clinical studies. The OGM technology is highly sensitive with an ability to detect all classes of clinically significant genome-wide SVs (aneuploidies, CNVs, balanced genomic rearrangements, repeat contraction, repeat expansions, and mosaicism)^12-14^. The use of OGM technology is of significant advantage in the analysis of POC tissue as it precludes the requirement for culturing and eliminates the risk of associated microbial contamination. However, MCC remains a challenge insignificant number of cases and impedes a definitive diagnosis. Although microarrays have been implemented as a reliable method of genome-wide analysis for pregnancy losses, and have shown increased diagnostic yield over conventional karyotyping^15-19^, they cannot circumvent a high percentage of MCC contamination. To address the significant variations in maternal and fetal content in the POC specimens, an additional pre-analytical step was added where the specimens were subjected to hematoxylin and eosin (H&E) histologic staining and examination by a pathologist to assess the presence of chorionic villi before any diagnostic test was performed on these specimens. The slides were further macro-dissected for the enrichment of Chorionic villi and then analyzed using SNP microarray methodology. Thus, in this brief report, we propose an integrated workflow that includes MCC characterization of POC tissue, followed by OGM for MCC negative cases or SNPM with histological assessment for MCC positive cases to improve the diagnostic yield in cytogenetic analysis of POC tissue.

## Materials and Methods

### Patient specimen

For six month period in 2021, nine POC cases were prospectively collected and analyzed using the proposed workflow along with conventional clinical testing (**Figure 1**). Additionally, we performed an internal audit at our institution to identify POC cases in the cytogenetics laboratory that were subjected to karyotype or SNPM analysis over eight years (2013-2021). Retrospectively, we identified and analyzed a total of 578 POC samples that were received, out of which only ∼6% of cases had clinically actionable information that contributed to patient care. Of these 578 POC samples, we re-evaluated 22 cases (for whom the FFPE blocks were available) on which no clinical diagnosis could be achieved with conventional cytogenetic analysis. Thus, the present study evaluated 31 POC specimens that included, 22 cases of spontaneous miscarriage and 9 cases of RPL. Clinical information was obtained from patient charts under an IRB-approved protocol, and archival FFPE blocks with slides were retrieved for review. The study was approved by the IRB A-BIOMEDICAL I (IRB REGISTRATION #00000150), Augusta University. HAC IRB # 611298. Based on the IRB approval, the need for consent was waived; all PHI was removed, and all data was anonymized before accessing for the study.

**Figure 1.**
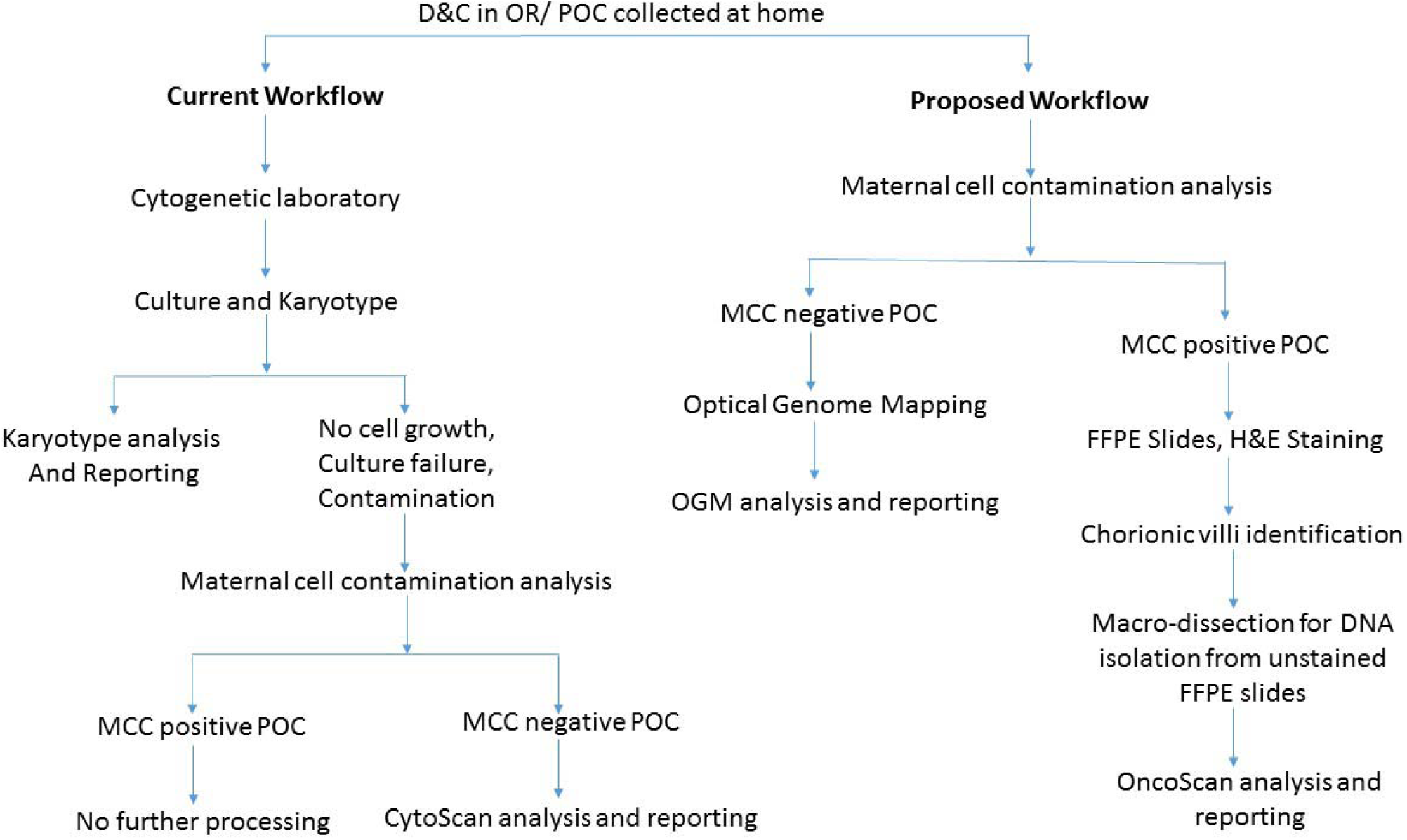
Flowchart depicting a modified diagnostic laboratory workflow compared to routine testing for high diagnostic yield in evaluating POC.

### Maternal cell Contamination

DNA samples from the mother’s blood and POC (fetal sample) were processed for maternal cell contamination analysis. Briefly, both samples were subjected to polymerase chain reaction (PCR) amplification of nine polymorphic STR loci plus amelogenin. Amplicons from the maternal and fetal samples were sized by capillary electrophoresis, and then directly compared to determine if maternal cell contamination was present in the fetal sample.

### Optical genome mapping

For POC negative for MCC, ∼15 mg of POC section was used to isolate ultra-high molecular weight (UHMW) using the Bionano Prep SP DNA Isolation Kit. Subsequently, the Bionano Prep DLS Labeling Kit was used to fluorescently label long molecules at specific sequence motifs throughout the genome. The labeled DNA was loaded onto Saphyr chips for linearization and imaging in massively parallel nanochannel arrays. The observed unique patterns on single long DNA molecules were used for de novo genome assembly and structural variant calling via the Bionano Solve pipeline (version 3.6).

### Histological assessment and SNPM analysis

FFPE slides were stained with H&E and examined by a board-certified pathologist (RK) to identify chorionic villi (fetal tissue) and marked for macrodissection (**Figures 2 and 3**). Following fetal tissue enrichment using macrodissection, DNA was isolated using a QIAamp DNA FFPE tissue kit (Qiagen, Germany). The isolated DNA was analyzed using the whole-genome SNP microarray following the manufacturer’s protocol (OncoScan® FFPE assay kit, ThermoFisher Scientific, USA). The platform consists of 220,000 markers throughout the entire genome. The test compares the samples to control samples from the HAPMap set of 270 individuals. The raw data was analyzed using the Chromosome Analysis Suite (ChAS) 4.0 software and were matched to *in silico* FFPE reference sets.

**Figure 2.**
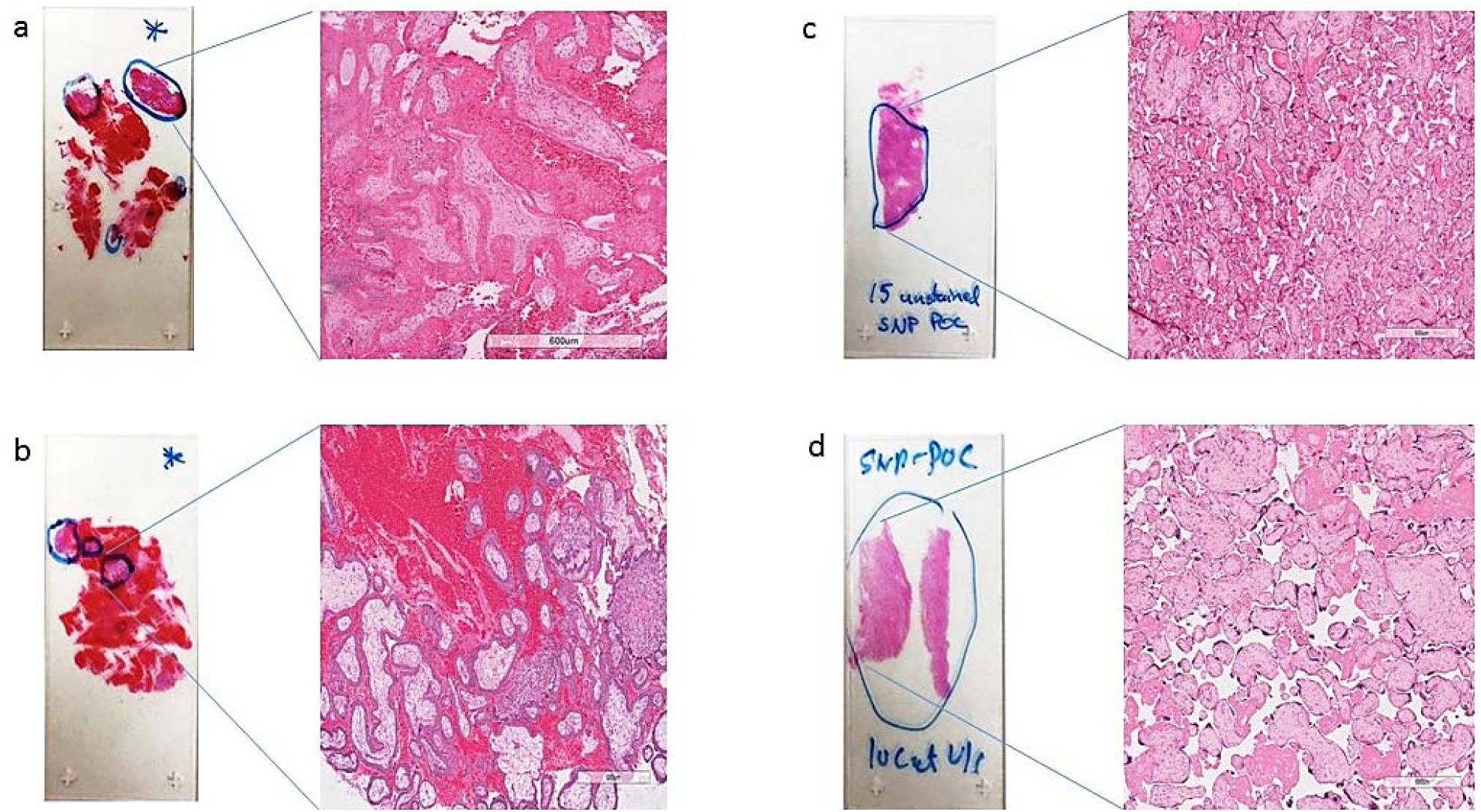
Representative H&E slides identifying (fetal tissue) with a zoomed-in view at 600 μm magnification, marked for macrodissection and DNA isolation by a surgical pathologist.

**Figure 3.**
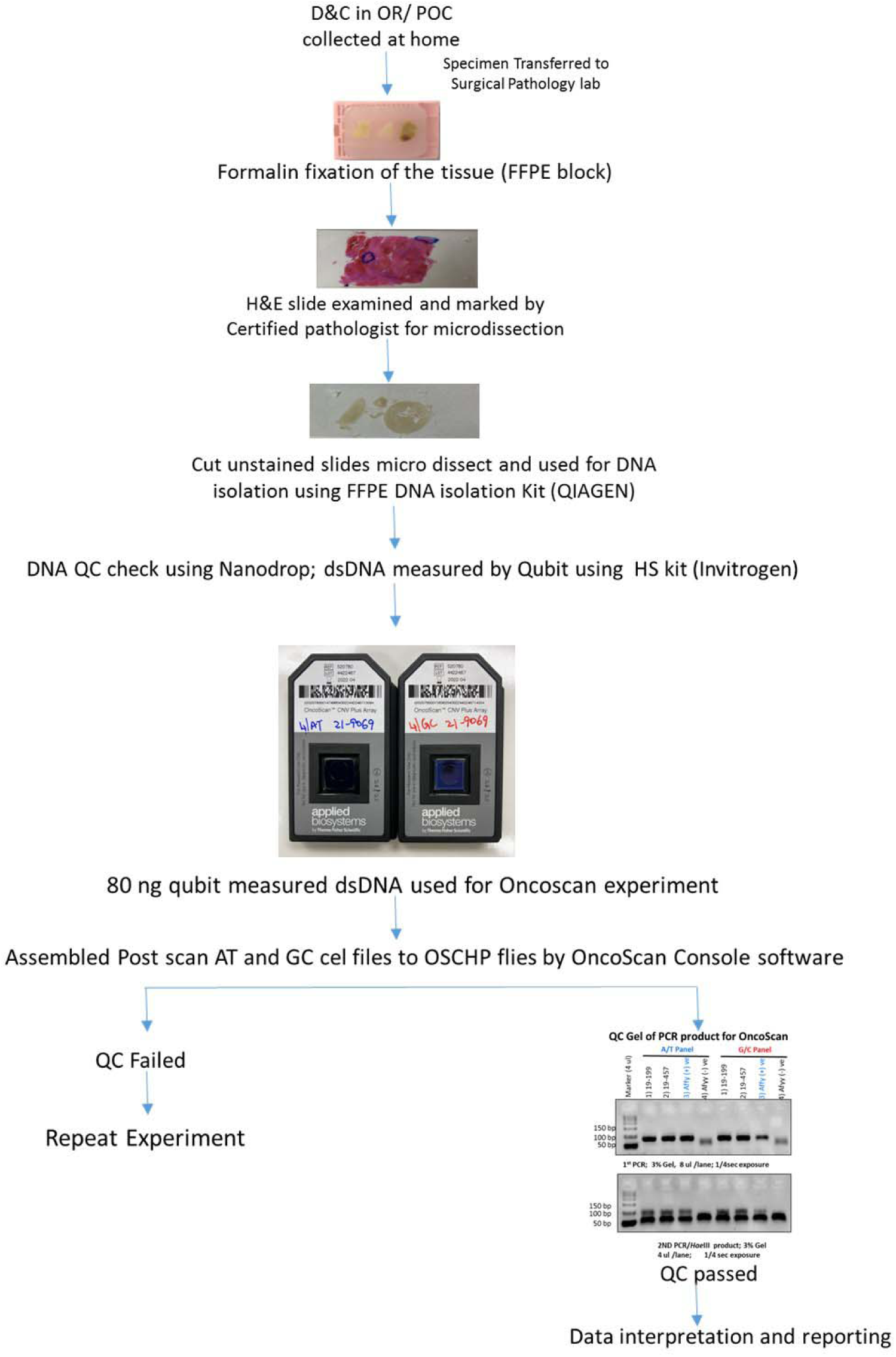
Flowchart depicting the detailed workflow for MCC positive POC tissue with histological assessment and Oncoscan microarray analysis and reporting.

## Data Analysis

The analysis results were classified and reported as follows: gains and losses of an entire chromosome as an aneuploidy, gains, and losses (>10 MB) of regions of a chromosome as partial aneuploidy, and terminal loss from one chromosome coupled with a terminal gain from another chromosome as an unbalanced translocation. The following databases were used to assess the clinical significance of genomic aberrations less than 5Mb: the database of genomic variants (DGV), DatabasE of genomiC varIation, and Phenotype in Humans using Ensembl Resources (DECIPHER), Online Mendelian Inheritance in Man (OMIM) and Pubmed. The American College of Medical Genetics and Genomics (ACMG) microarray reporting guidelines were used to classify the variants as pathogenic, likely pathogenic, and variants of uncertain clinical significance. Notably, benign and likely benign variants were not reported.

## Results

### Patient characteristics

In the present study, 31 POC samples were evaluated of which 28 did not yield clinical diagnostic results with conventional cytogenetics. The mean age of women was 33.5 ± 5.1 (range 22 – 44 years), with 22 cases of spontaneous miscarriage and 9 of RPL. Women with RPL had a total of 34 losses. Sixteen women had a history of no prior live births, while eight, four, and three women had one, two, and three prior births, respectively. Of these cases, twenty-seven women had a miscarriage in < 20 weeks of gestation, three had intra-uterine fetal demise, and one had a stillbirth (**Table 1**).

**Table 1.** Clinical characteristics.

| Patient characteristic | Classification | No. |
| --- | --- | --- |
| Age (Mean ± SD) | | 33.4 ± 5.1 |
| Sporadic miscarriage |  | 22 |
| Recurrent pregnancy loss |  | 9 |
| No. of losses | 2 | 4 |
|  | 3 | 4 |
|  | 4 | 0 |
|  | 5 | 1 |
|  | 6 | 1 |
|  | 7 | 1 |
| No. of live births | 0 | 16 |
|  | 1 | 8 |
|  | 2 | 4 |
|  | 3 | 3 |
| Miscarriage | <20 weeks | 27 |
|  | Intra-uterine fetal demise | 3 |
|  | Stillbirths | 1 |

### Maternal cell Contamination

Of the nine prospectively collected POC cases, six were identified with MCC contamination and were subjected to histological assessment and SNPM analysis. The remaining three cases were processed for OGM analysis. Of these three cases, two cases were analyzed using karyotyping, while the cells could not be grown for the third case, and were analyzed using microarray analysis as per the current clinical cytogenetic workplan.

### Optical genome mapping analysis

The three samples analyzed using OGM achieved excellent quality control performance metrics with an average N50 of 292 kbp (recommended >220 kbp), map rate of 87% (recommended >70), and label density of 15.8 (recommended 15-17). OGM was concordant in identifying the results observed with conventional cytogenetic analysis. OGM confirmed trisomy 15, and 21 in the first two cases, previously identified by karyotyping, while in the third case no reportable SVs or CNVs were detected using OGM, as previously reported with microarray technology (**Figure 4**).

**Figure 4.**
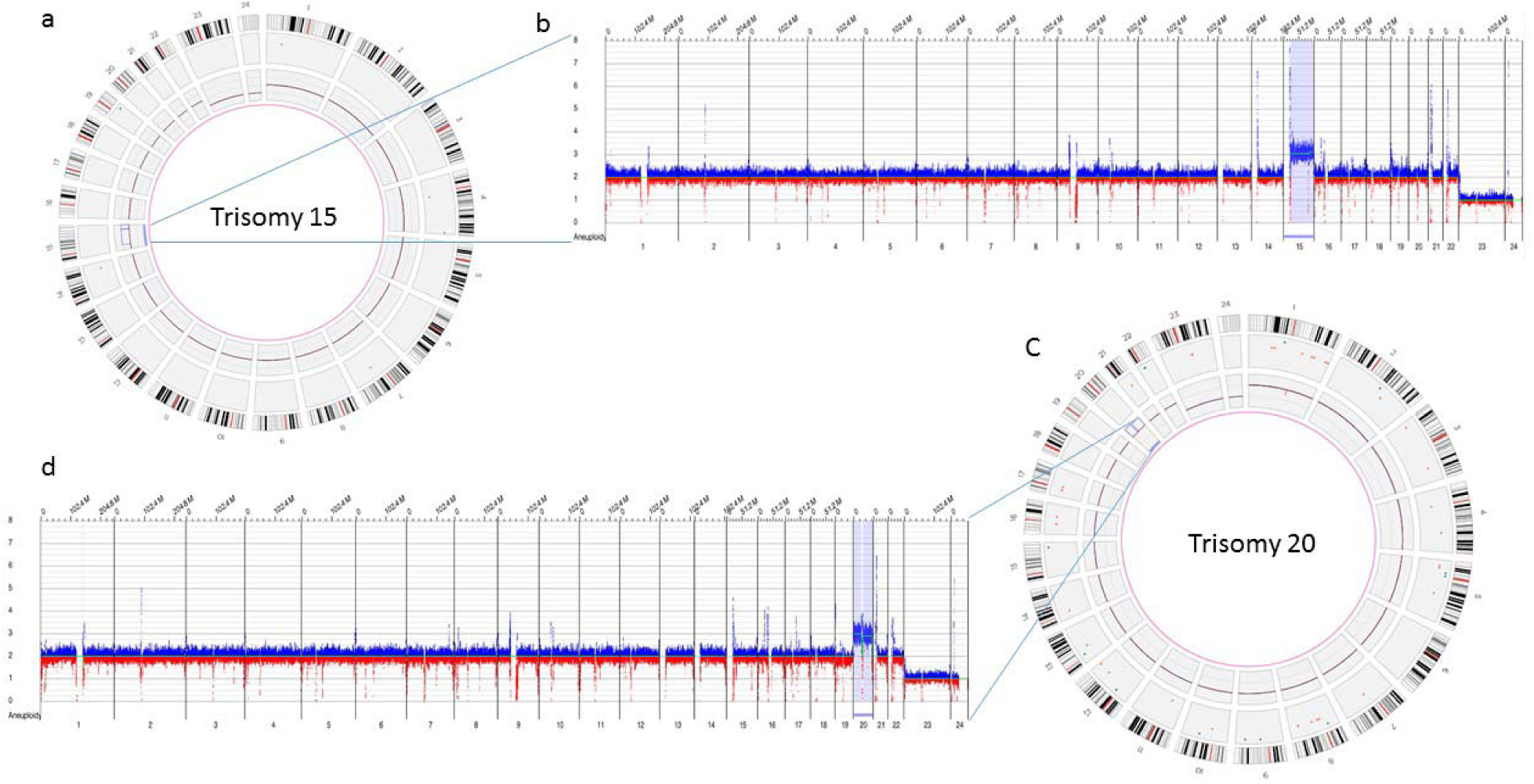
Optical genome mapping identifying trisomy 15 a) circus plot showing trisomy 15 b) copy number track view with trisomy 15, c) circus plot showing trisomy 20 b) copy number track view with trisomy 20.

### Histological assessment and SNPM analysis

Of the twenty-eight FFPE POC samples macro-dissected for Chorionic villi and analyzed utilizing the OncoScan assay, results were obtained in 92.8% (26/28) cases, where traditional cytogenetic analysis had not yielded a definitive result. Maternal cell contamination was identified in 7.1% (2/28) cases, wherein copy number changes could not be identified; however, both cases were identified as males. Genetic aberrations were detected in 42.8% (12/28) cases, while no reportable copy number aberrations or absence of heterozygosity (AOH) were detected in 50.0% (16/26) cases. (**Table 2**).

**Table 2.** Abnormal genetic aberrations detected with SNP microarray and optical genome maping on the product of conception.

| S.No. | Trimester | ISCN Nomenclature | Size |
| --- | --- | --- | --- |
| **OncoScan Analysis** |  |  |  |
| 1 | 9 weeks, 1st | arr[hg19] (22)x3 | Entire chromosome 22 |
| 2 | 10 weeks, 1st | arr[hg19] 4q34.3(178,112,003-182,153,124)x3, (21,22)x2~3 | 4.0 Mb Gain of chr 4 and Gain of entire chromosomes 21 and 22 |
| 3 | 8 weeks, 1st | arr[hg19] (8)x2~3 | Entire chromosome 8 |
| 4 | 9 weeks, 1st | arr[hg19] (8)x3 | 146.1Mb |
| 5 | 10 weeks, 1st | arr[hg19] (14)x2~3 | Entire chromosome 14 |
| 6 | 10 weeks, 1st | arr[hg19] (X)x1~2 | Entire chromosome X |
| 7 | 14 weeks, 2nd | arr[hg19]2q34q37.2(214027641-236628038)x3, 2q37.2q37.3(236228142-243052331)x1 | 22.6Mb gain and 6.4Mb loss |
| 8 | 11 weeks, 1st | arr [hg19] (1-21,X)x2, (22)x2~3 | Entire chromosome 22 |
| 9 | 8 weeks 5 days, 1st | arr[hg19] (15)x2~3 | Entire chromosome 15 |
| 10 | 6 weeks, 6 days, 1st | arr[hg19] (18)x3 | Entire chromosome 18 |
| 11 | 26 weeks 1 day, 2nd | arr[hg19] 2p21(43411752-44710936)x3, 21q21.1(18762223-19136546)x3 | 1.29 Mb Gain and 374 Kb Loss |
| 12 | Less than 3 weeks, 1st | arr[hg19] 13q14.11(42311546_42413745)x1 | 102 Kb Loss |
| **Optical Genome Mapping** |  |  |  |
| 13 | 6 weeks, 1st | ogm[GrCh38](15)x3 | Entire chromosome 15 |
| 14 | 8 weeks, 1st | ogm[GrCh38](20)x3 | Entire chromosome 20 |

### Proposed workflow compared to conventional cytogenetic testing

In the proposed workflow, a result was obtained in 93% (29/31) of the cases compared to 9.6% (3/31) with routine workflow, while the diagnostic yield was 45.1% (14/31) with the proposed workflow compared to 6.4% (2/31) with conventional cytogenetic testing.

### Selected, Interesting Clinical cases

A female (>35 year age) with a history of RPL and multiple failed attempts at karyotyping had a miscarriage in the 1^st^ trimester in her eighth pregnancy. The patient had a medical history of Hashimoto’s thyroiditis, asthma, obesity, insulin resistance, Cushing disease, and depression. H&E stained slides of FFPE tissue were marked by a surgical pathologist for chorionic villi which were macro-dissected for DNA isolation. OncoScan analysis identified double mosaic trisomies involving chromosomes 21 and 22 (ascertained as ∼70% mosaic gain), and a copy number gain of Chr 4q, a variant of uncertain significance (**Figure 5**).

**Figure 5.**
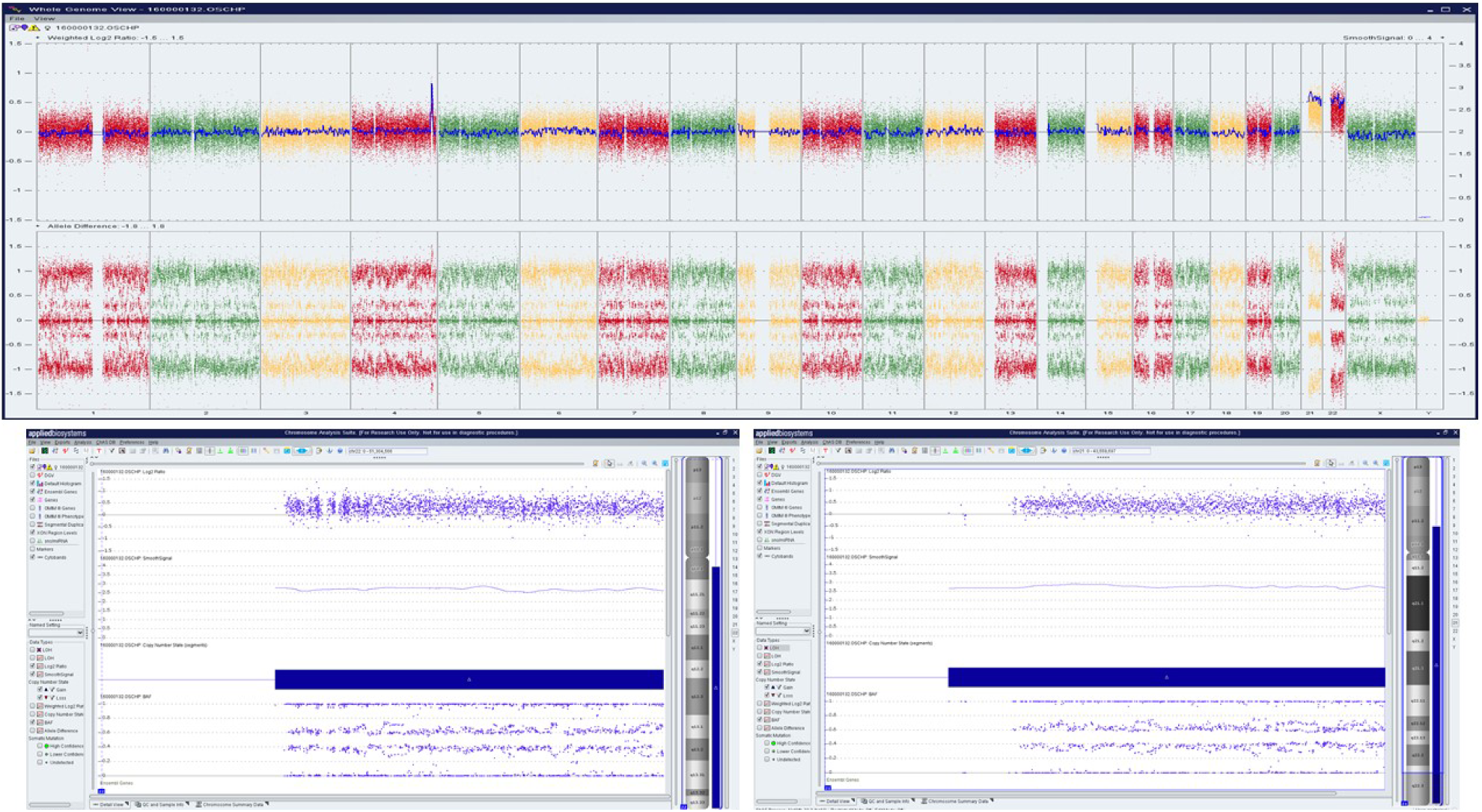
OncoScan analysis identifying double mosaic trisomies involving chromosomes 21 and 22 (ascertained as ∼70% mosaic gain), and a copy number gain of Chr 4q, a variant of uncertain significance: a) Whole-genome view, b) log2ratio, smooth signal, and BAF of chr 21, c) log2ratio, smooth signal, and BAF of chr 22

A female (>30 year age) with a history of RPL and multiple failed attempts at karyotyping had a miscarriage in the 2^nd^ trimester of her sixth pregnancy. The patient had a medical history of hypothyroidism, recurrent pregnancy loss and was reported to harbor a compound heterozygous *MTHFR* gene mutation. Cytogenetic/karyotyping could not be performed on this specimen, as no metaphase cells were present. On analyzing the FFPE POC tissue with macrodissection, a 22.6 Mb copy gain of Chr 2q34q37.2 and a 6.4 Mb copy loss of Chr 2q37.2q37.3 was identified, indicating a complex rearrangement involving the long arm of chromosome 2 (**Figure 6**).

**Figure 6.**
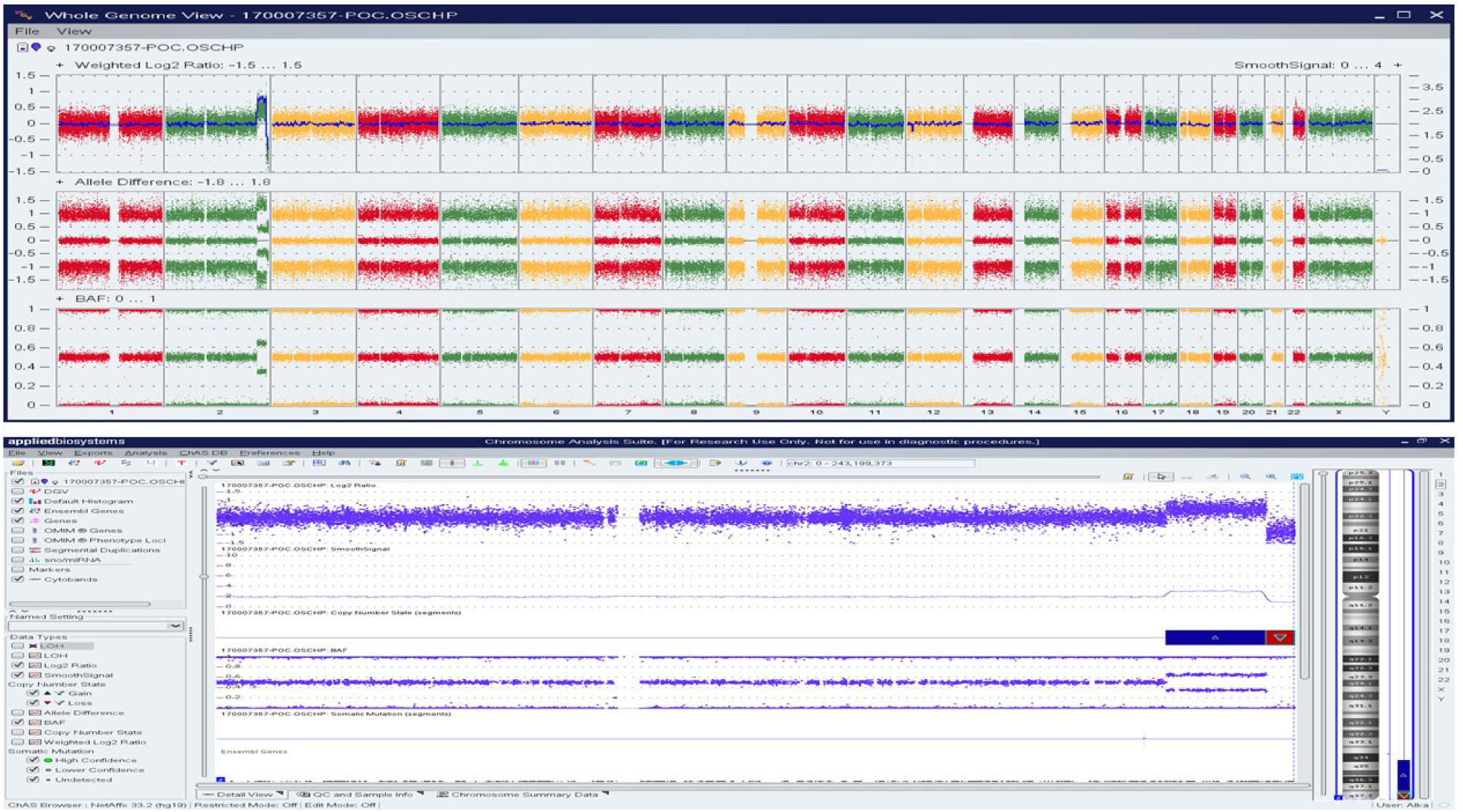
OncoScan analysis identified 22.6 Mb copy gain of Chr 2q34q37.2 and a 6.4 Mb copy loss of Chr 2q37.2q37.3, indicating a complex rearrangement involving the long arm of chromosome 2: a) whole-genome view, b)) log2ratio, smooth signal, and BAF of chr 2.

## Discussion

Pregnancy losses occur in ∼15-25% of clinically recognized pregnancies, with approximately 5-10% caused by chromosomal abnormalities. Identification of these genetic aberrations is essential in understanding the underlying cause of these painful and traumatic events. Defining the genetic etiology of these pregnancy losses allows these patients to understand the reason for their miscarriage, bringing some degree of closure, and may also predict the recurrence risk for the parents.^1,4,5^ The chromosomal abnormalities in the POC tissue can also suggest further testing protocols, with certain abnormalities such as aneuploidies that have not been associated with recurrence-risk and may not require further testing, while unbalanced chromosomal translocation or inversion and euploid POC are suggestive indications for further testing including whole-genome chromosome analysis of parents, and RPL workup, respectively.^20^ However, performing chromosome analysis on POC tissue is not always feasible and a diagnosis is precluded because of a variety of reasons. Conventional cytogenetic analysis is limited because of the requirement of live cells for culturing, culture contamination, and maternal cell contamination. Further, conventional techniques are limited in resolution or biased to certain loci and thus, several genetic events may remain cryptic. The low diagnostic yield observed with conventional cytogenetic technique/workflow led us to devise an integrated workflow that bypasses the requirement for culturing (a major time-consuming step). The proposed workflow begins with MCC characterization of POC tissue, followed by OGM for MCC negative cases or SNPM with histological assessment for MCC positive cases.

Optical genome mapping is an emerging cytogenetic technology that can detect all classes of SVs and CNVs in a single assay as compared to conventional cytogenetic techniques (karyotyping, FISH and SNPM). Considering OGM to be the most sensitive and high-resolution cytogenetic technology, the primary aim was to assess the feasibility of performing OGM for the analysis of the POC tissue. Although only three samples were analyzed using OGM, the excellent quality control metric demonstrates the feasibility and utility of using OGM for the analysis of POC tissue. Several of the POC tissues remain uncharacterized because of failed cultures which can be adequately addressed by OGM technology as UHMW DNA was isolated directly from the POC tissue. Further, the use of OGM reduces the time to reach a diagnosis as cultures may take weeks before sufficient cells are available for analysis. As OGM technology has demonstrated the ability to identify additional/novel structural variants that were missed by conventional methods in several phenotypes,^12-14^ it would be interesting to see if OGM would detect additional aberrations in otherwise normal POC genomes evaluated with conventional techniques.

For the MCC positive cases, the addition of a pre-analytical step with histological assessment and SNPM analysis led to a definitive result in 90.5% (19/21) samples. The two samples that still showed maternal cell contamination, were identified as males, although a copy number assessment could not be made. It is important to note that maternal cell contamination as low as 5-10% is detectable with SNP microarray, and insignificant levels of MCC have been reported in 93% of fresh and 60% of FFPE tissue. However, an MCC of less than 50% may not severely affect the detection of segmental or whole chromosome imbalances.^17^ The high diagnostic yield in this study is partially attributed to the histological assessment of the FFPE POC tissue for Chorionic villi, which seems to be an ideal methodology upstream of SNP microarray analysis. The histological assessment helps rule out the presence of maternal tissue and enables macrodissection of Chorionic villi, to enrich the fetal tissue for DNA isolation and SNP microarray analysis. The major limitation that precludes a definitive clinical diagnosis with SNP microarrays is the issue of MCC,^20^ which can be addressed with this approach in the pre-analytical stage of the assay. The ability of the platform to yield a diagnosis with minimal MCC should therefore be more fully exploited for clinical insights, and the histological assessment of the FFPE POC tissue might be a necessary additional step that needs to be incorporated in the clinical workup for improved diagnosis. The higher performance and the clinical utility of the SNP microarray were further highlighted by the two RPL cases where multiple attempts were made to obtain a karyotype analysis but no clinical diagnosis could be achieved, and a definitive clinical diagnosis with genetic aberrations was detected on analyzing the FFPE POC tissue.

Overall, we propose an integrated workflow with these technologies that demonstrated the clinical utility and higher diagnostic yield in evaluating POC specimens in this study. The OGM technology is highly sensitive to detect several classes of structural variants and copy number variants, while SNPM with the addition of this pre-analytical step has a significant potential to improve clinical diagnosis in this important area of reproductive medicine. Although, the study is limited to a small sample size, the critical insights obtained with this distinct approach address the key issues associated with current diagnostic testing, and might serve as an important factor to improve diagnostic yield in POC analysis.

## Data Availability

All relevant data in the present work is contained in the manuscript.

## Ethics approval

The study was approved by the IRB A-BIOMEDICAL I (IRB REGISTRATION #00000150), Augusta University. HAC IRB # 611298. Based on the IRB approval, the need for consent was waived; all PHI was removed, and all data was anonymized before accessing for the study.

## Declarations

RK has received honoraria, and/or travel funding, and/or research support from Illumina, Asuragen, QIAGEN, Perkin Elmer Inc, Bionano Genomics, Agena, Agendia, PGDx, Thermo Fisher Scientific, Cepheid, and BMS. AH, and AC are salaried employee at Bionano Genomics Inc. All other authors have no competing interests to disclose.

